# Phase-Amplitude Coupling in Autism Spectrum Disorder: Results from the Autism Biomarkers Consortium for Clinical Trials

**DOI:** 10.1101/2022.09.25.22279830

**Authors:** Fleming Peck, Adam J. Naples, Sara J. Webb, Raphael A. Bernier, Katarzyna Chawarska, Geraldine Dawson, Susan Faja, Shafali Jeste, Michael Murias, Charles A. Nelson, Frederick Shic, Catherine Sugar, Damla Şentürk, James C. McPartland, April R. Levin, the ABC-CT Network

**Affiliations:** Department of Neurology, Boston Children’s Hospital, Harvard Medical School; Department of Psychology, University of California, Los Angeles; Child Study Center, School of Medicine, Yale University; Center for Child Health, Behavior and Development, Seattle Children’s Research Institute; Department of Psychiatry and Behavioral Sciences, University of Washington; Duke Institute for Brain Sciences, Duke University; Duke Center for Autism and Brain Development, Duke University; Department of Psychiatry and Behavioral Sciences, Duke University; Laboratories of Cognitive Neuroscience, Division of Developmental Medicine, Boston Children’s Hospital, Harvard Medical School; Department of Pediatrics, Children’s Hospital Los Angeles, University of Southern California Keck School of Medicine; Institute for Innovations in Developmental Sciences, Northwestern University; Department of Pediatrics, University of Washington; Department of Biostatistics, University of California, Los Angeles; Department of Psychiatry and Biobehavioral Sciences, University of California, Los Angeles

**Author notes:** Corresponding Authors: April R. Levin, James C. McPartland.

## Abstract

**Background:** Autism spectrum disorder (ASD) is defined behaviorally, but measures that probe underlying neural mechanisms may provide clues to biomarker discovery and brain-based patient stratification with clinical utility. Phase-amplitude coupling (PAC) has been posited as a measure of the balance between top-down and bottom-up processing in cortex, as well as a marker for sensory processing and predictive coding difficulties in ASD. We evaluate differences in PAC metrics of resting-state brain dynamics between children with and without ASD and relate PAC measures to age and behavioral assessments.

**Methods:** We analyzed electroencephalography data collected by the Autism Biomarkers Consortium for Clinical Trials, including 225 (192 male) ASD and 116 (81 male) typically-developing children aged 6-11 years. We evaluated the strength and phase preference of PAC and the test-retest reliability of PAC across sessions.

**Results:** There was significantly increased alpha-gamma and theta-gamma PAC strength in ASD. When considering all participants together, we found significant associations of whole brain theta-gamma PAC strength with measures of social communication (Beta = 0.185; p = 0.006) and repetitive behaviors (Beta = 0.166; p = 0.009) as well as age (Beta = 0.233; p < 0.0001); however, these associations did not persist when considering the ASD group alone. There are also group differences in theta-gamma phase preference.

**Conclusions:** This large, rigorously collected sample indicated altered PAC strength and phase bias in ASD. These findings suggest opportunities for back-translation into animal models as well as clinical potential for stratification of brain-based subgroups in ASD.

## Introduction

Autism spectrum disorder (ASD) is characterized by challenges with social communication, repetitive behaviors, and sensory processing. A core feature of neural circuit difference in ASD appears to be homeostatic imbalance at the level of neural circuits that can occur as a result of multiple underlying differences in physiological mechanisms; by various accounts, this has been described as an imbalance in feedforward versus feedback processing (1,2), excitation versus inhibition (3,4), short-range versus long-range connectivity (5), or signal versus noise (6,7). This macroscale circuit activity can be sampled noninvasively using electroencephalography (EEG).

Many of the most common EEG analyses in ASD have focused on resting-state spectral power, with some studies showing increased power in higher frequencies (8–10) and/or decreased power in mid-range frequencies (11,12). Importantly, while these spectral power analyses capture the overall strength of oscillations averaged over an entire EEG recording session, the process of balancing neural circuit activity is dynamic rather than static (14). Levels of excitation and inhibition, for example, do not remain stationary over the course of minutes in an active circuit. Instead, they actively adjust, on the order of milliseconds, in response to the ongoing activity of the circuit and its neighbors. Therefore, EEG analyses that capture not only the *amount* of activity, but also the *timing* thereof, may provide further insights into the neural circuit activity occurring in ASD.

Cross-frequency coupling has emerged as a method of analyzing this dynamic, moment-to-moment interaction between oscillations of different frequencies within EEG signal. Specifically, phase-amplitude coupling (PAC) evaluates how the phase of low frequency oscillations modulates the amplitude of high frequency activity. Strong coupling acts as a clocking mechanism in the brain, creating perceptual windows that integrate and segregate temporally relevant and irrelevant information, respectively (15). The phase and frequency of PAC differs by cortical layer (16,17) and is thus postulated to reflect the direction (bottom-up versus top-down) of information flow in the cortex (17–19). The balance between top-down and bottom-up processing in cortex may serve as a marker for sensory processing and predictive coding (17,20), which are of particular relevance to ASD given theories of sensory prediction impairment (21) and disruption of executive function development (20). Importantly, PAC findings can be translated from analyses in cortical organoids (22) and animal models (16,17,23,24), and from a clinical standpoint, PAC offers potential for measuring treatment response via manipulation by medication (25) and neuromodulation (26).

Previous resting-state PAC analyses based on MEG source-space data in ASD have focused on alpha-gamma coupling with findings of increased parietal-occipital coupling (28) and increased central and decreased lateral coupling (29). In addition, PAC has been shown to be altered in ASD compared to controls during visual processing tasks (30), including face processing (31–33). In individuals with Phelan-McDermid Syndrome (a neurogenetic disorder with high prevalence of ASD), the phase bias of alpha-gamma PAC on EEG sensor-space data is altered most particularly in posterior electrodes, and the magnitude of PAC is associated with restricted and repetitive behaviors (19). In the face of this prior work consisting of EEG and MEG brain recordings, resting-and task-related paradigms, and study of ASD of neurogenetic or idiopathic etiology, resting-state PAC in idiopathic ASD has yet to be analyzed with EEG.

The Autism Biomarkers Consortium for Clinical Trials (ABC-CT) provides the opportunity to evaluate resting-state EEG PAC in a large, multisite, longitudinal sample with rigorously collected data using scalable technologies (34). The ABC-CT was designed to facilitate biomarker development by collecting neuroimaging and behavioral data from a large sample of ASD and typically-developing (TD) participants to evaluate individual differences of objective measures. The main goal of the present manuscript is to evaluate PAC metrics that are characteristic of brain function in children with ASD as a step towards developing clinically relevant biomarkers.

## Methods and Materials

### Participants

This study drew from data collected for the ABC-CT (34). Children with genetic or neurologic disorders causally related to ASD were excluded from the study, and medication was allowed given a stable regimen of at least 8 weeks preceding enrollment. Inclusion criteria included IQ between 60 and 150 in the ASD group, or between 80 and 150 in the TD group, as measured by the Differential Ability Scales, Second Edition. Data were collected longitudinally from a total enrollment of 280 ASD and 119 TD participants aged 6-11 years old across 5 different sites (Boston Children’s Hospital; University of California, Los Angeles; University of Washington; Duke University; and Yale University). At the first (baseline) timepoint, 211 ASD and 106 TD (317 total) participants contributed adequate EEG data for inclusion. At the second time point collected 6 weeks later, 215 ASD and 100 TD (315 total) participants provided EEG data. 174 ASD and 88 TD (262 total) participants contributed data at both timepoints. Demographic information for participants who contributed adequate EEG data at either timepoint is included in Table 1.

**Table 1:** Demographic information by diagnostic group.

|  | ASD (n = 252) | TD (n = 116) | Statistical test |
| --- | --- | --- | --- |
| Sex (n) | 192 M/60 F | 81 M/36 F | $\chi^2(1) = 1.67, p = .20$ |
| Age (years) (mean ± std (range)) | 8.6 ± 1.6 (5.5) | 8.5 ± 1.6 (5.5) | $F(1,366) = 0.99, p = 0.95$ |
| Race (%) | | | $\chi^2(4) = 9.05, p = .06$ |
| Asian | 6 | 1.7 |  |
| Black | 7.5 | 3.4 |  |
| Mixed race | 15.5 | 12 |  |
| White | 68.3 | 82.1 |  |
| Other | 2.4 | 0.9 |  |
| Ethnicity (Hispanic or Latinx) | 17 | 6 | $\chi^2(1) = 7.46, p = .006$ |
| Behavioral assessments (mean ± std (range)) | | | |
| ADOS Total Scaled Score | 14.2 ± 4.7 (18) | 2.4 ± 1.8 (9) | $F(1,366) = 5.25, p < .001$ |
| Full Scale IQ | 97.6 ± 17.8 (90) | 115.3 ± 12.4 (63) | $F(1,366) = 2.07, p < .001$ |

A central Institutional Review Board at Yale University approved the study protocols. Written informed consent was obtained from a parent or legal guardian and assent was obtained from each child before their participation in the study.

### Paradigm

Participants watched six soundless videos akin to screensavers. Each video played forward for 15 seconds and then in reverse for 15 seconds. The six videos were played in three blocks of two videos each at 30 frames per second. Video display was limited to 7 × 9.3 cm to minimize eye movement during task completion.

### Data collection

EEG data were collected in full lighting. A behavioral assistant facilitated data collection by directing the child participant to pay attention to the screen. Placement of behavioral assistant differed across sites (but was standardized within sites). Data were collected with 128-channel Hydrocel Geodesic Nets and NetStation software with a sampling rate of 1000 Hz.

### EEG data processing

Data were processed using the Batch EEG Automated Processing Platform (BEAPP) (35), which allows all EEG files to be processed with the same artifact removal criteria, via the Harvard Automated Processing Pipeline for EEG (HAPPE) (36). The data were filtered using a 1 Hz digital high-pass filter and 100 Hz low-pass filter and then resampled to 250 Hz. A spatially distributed subset of 18 channels (10-20 montage) were selected for further preprocessing via HAPPE and analysis. The artifact removal process consisted of 60 Hz electrical noise removal via CleanLine’s multitaper method (37), bad channel rejection, and movement, muscle, and eye blink artifact removal with wavelet-enhanced independent component analysis (ICA) and ICA with Multiple Artifact Rejection Algorithm (MARA) (38). Bad channels were repopulated using spherical interpolation, and data were re-referenced to average. Each of the 30-second videos allowed for three non-overlapping, continuous 10-second EEG data segments, and a 2-second buffer was included at the beginning and end of each EEG segment as a buffer to avoid edge effects from filters (Figure 1B). Segments were rejected based on HAPPE’s amplitude rejection criteria.

**Figure 1:**
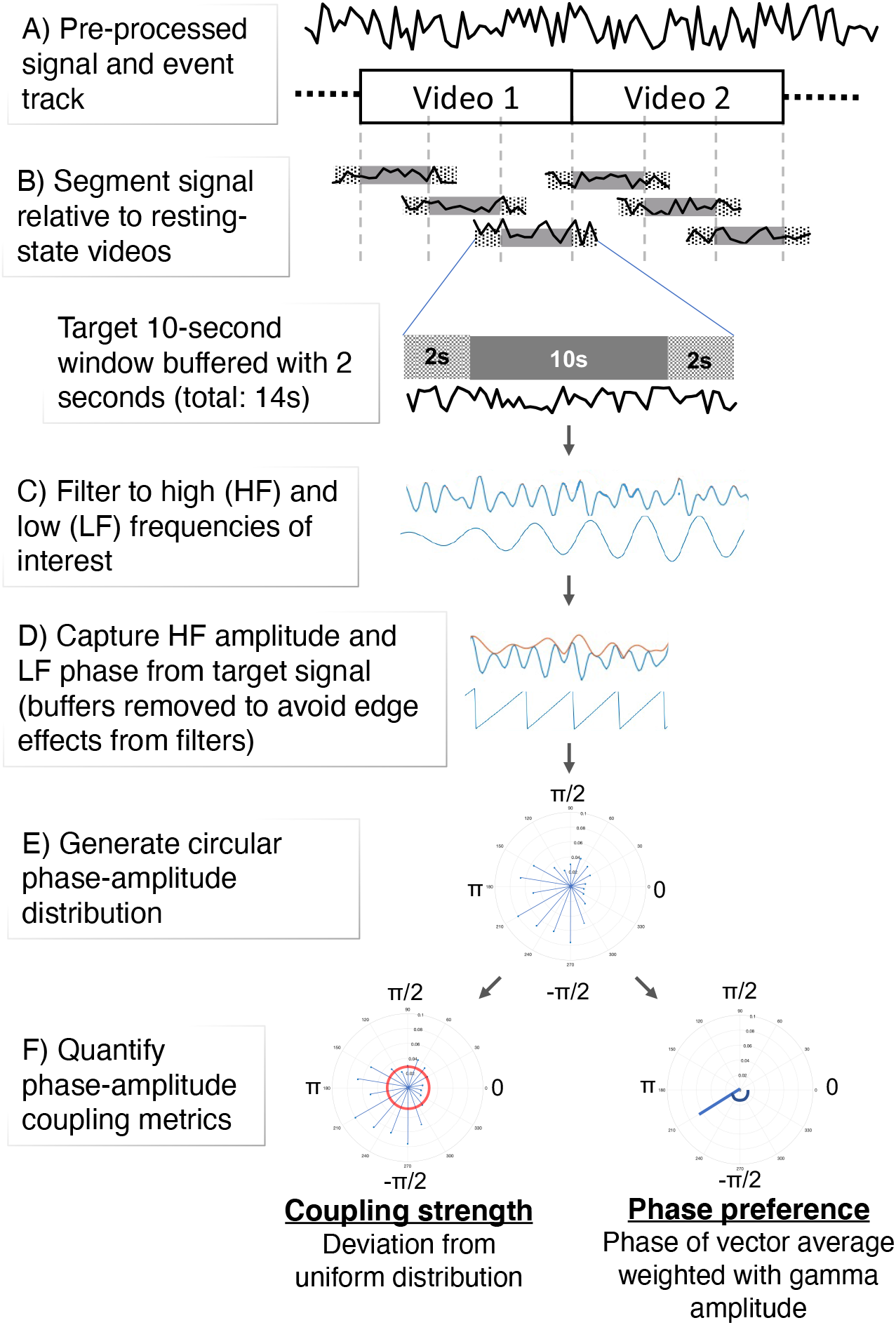
Visualization of phase-amplitude coupling analysis steps.

### EEG rejection criteria

EEG files that were within three standard deviations of the following HAPPE data quality metrics thresholds were included in analysis: retained variance (>7.5), percentage of good channels (>78%), percentage of independent components rejected (<80%), and mean and median retained artifact probability (<0.23, <0.18, respectively). 66 (16.5%) and 84 (21%) files were excluded at the first and second timepoints, respectively. HAPPE data quality metrics for each group are listed in Table 2.

**Table 2:** HAPPE data quality metrics by diagnostic group. **Student’s t-test p < 0.001; * Student’s t-test p < .005 (ASD vs. TD)

|  | Timepoint 1 |  | Timepoint 2 |  |
| --- | --- | --- | --- | --- |
|  | ASD (n = 211) | TD (n = 106) | ASD (n = 215) | TD (n = 100) |
| HAPPE metrics (mean ± std (range)) | | | | |
| Independent components rejected (%) | 33.6 ± 14.3 (70.9)** | 25.9 ± 11.8 (50.7) | 0.32 ± 0.1 (0.61)** | 0.25 ± 0.1 (0.5) |
| Variance retained (%) | 68.3 ± 19.7 (92.1)* | 75.1 ± 17.5 (87.9) | 70.5 ± 16.7 (72.2)** | 78.4 ± 15.8 (69.5) |
| Good channels (%) | 96.5 ± 4.8 (16.7) | 96.4 ± 5.4 (16.7) | 96.4 ± 4.9 (16.7) | 96.1 ± 5.1 (16.7) |
| Mean remaining artifact probability | 0.07 ± 0.04 (0.19) | 0.07 ± 0.04 (0.2) | 0.08 ± 0.04 (0.19)* | 0.07 ± 0.04 (0.15) |

### Phase-amplitude coupling

The following PAC analysis steps were computed using the *pactools* Python toolbox (39). First, narrowband neural oscillations at low (2-20 Hz in 2 Hz steps) and high (20-100 Hz in 4 Hz steps) frequencies were extracted (Figure 1C). PAC was evaluated in each of these 210 low (LF) and high (HF) frequency pairs for all 18 channels. LFs were filtered with a 2 Hz bandwidth around the target low frequency, and HFs were filtered to 2 Hz below the target frequency with a variable upper bandwidth equal to the low frequency of interest. This filtering technique has shown to be preferable to static filtering for high frequencies (28). The amplitude of the HF and phase of the LF at each sample in time were extracted as described in (38) (Figure 1D). The phase of the LF time series was considered in 18 bins of width 20 degrees ranging -180° to 180°. and the HF amplitudes were binned according to the corresponding low frequency phase in which they occurred (Figure 1E). Then, the amplitude values in each bin were averaged, and the entire distribution was normalized by dividing each bin average by the sum of all bin averages. This process was repeated for 6 segments per participant (60 seconds of data total), and the resulting phase-amplitude distributions from the segments were averaged within a participant to minimize noise.

### Phase-amplitude coupling strength

Matlab R2019b was used to compute all PAC metrics and higher-level analyses unless otherwise noted. Coupling strength, or modulation index (MI), was computed as the Kullback-Leibler divergence of the phase-amplitude distribution from a uniform distribution (40) (Figure 1F). To isolate the effect of phase-amplitude coupling (from, for example, related factors such as spectral power), a surrogate distribution that did not represent real, time-locked effects of PAC was generated by offsetting the HF and LF signals by a randomly selected duration between 0.1 to 1.9 seconds and then repeating the phase-amplitude coupling analysis methods. The surrogate distribution was composed of 200 iterations of this null PAC strength calculation process, which was then averaged to obtain a surrogate modulation index and used to compute a z-scored PAC strength index of the original PAC computation (method proposed in (39)). Regional PAC strength was computed by averaging the normalized PAC strength over all frequency pairings of interest.

While the normalized modulation index quantifies the strength of phase-amplitude coupling, the phase preference measure represents the phase at which coupling primarily occurs. The preferred phase was determined as the phase of the average of vectors with angle of each LF phase bin weighted with corresponding HF amplitude (computed using the circ_mean function in the Circular Statistics Toolbox (41) (Figure 1F)).

### PAC Strength: A priori defined regions of interest and canonical wavebands

We primarily evaluated PAC metrics across three brain regions: the anterior (FP1, FP2, F3, F4, F7, F8, Fz), posterior (O1, O2, T5, T6, Pz), and whole (all 10-20 channels) brain. These regions were chosen based on prior studies showing that PAC often differs in anterior versus posterior channels (18,19). PAC was evaluated in three specific frequency range pairs following the canonical wavebands: theta (4-6 Hz) and gamma (28-56 Hz), alpha (8-12 Hz) and gamma (28-56 Hz), and low beta (14-20 Hz) and gamma (44-56 Hz). The lower end of the gamma range was increased when paired with low beta to avoid overlap in frequencies included with HF and LF signal filtering.

### PAC Strength: Data-driven grouping analyses

We also evaluated data-driven tests of PAC by comparing coupling strength between groups in frequency- and channel-space regions of significant PAC. First, we identified regions of significant PAC in frequency space within each of 18 channels for the ASD and TD groups separately. We employed the clustering procedure detailed in (18). In summary, a t-test was used to compare the raw MI values and the surrogate MI values (each an average of 200 iterations) from all participants of a diagnostic group. A “frequency grouping” comprised of adjacent significant frequency pairs without considering diagonal connections within a channel. To determine the significance of frequency groupings, over 1,000 iterations, the raw and surrogate MI values were flipped in a randomly selected subset of *n* participants ranging from 1 to *n*, and the test statistics and frequency grouping sizes were computed. Frequency groupings that were smaller than the 95^th^ percentile of this null frequency grouping size distribution were excluded.

### Phase preference analyses

Phase preference was computed as the vector average of the phase-amplitude distribution. The vectors were defined with the middle degree of a phase bin with magnitude of the average HF amplitude of the bin and were averaged using the circ_mean *Matlab* function (42). Group differences were statistically evaluated with the Watson-Wheeler test (effectively a t-test adapted for circular statistics) using the watson.wheeler.test *R* function.

We also evaluated the proportion of participants that expressed the highest HF amplitude in each LF phase (referred to as *max phase preference*). For each participant, at each channel and frequency pair of interest, we determined the LF phase bin with maximum HF amplitude. We then determined the number of participants that had their maximum HF amplitude in each LF phase bin and divided by the total number of participants, thus computing the proportion that each LF phase corresponded with the highest HF amplitude within a frequency- and channel-space region. We used t-tests to statistically compare the max phase bin proportions between diagnostic groups, and significant differences were determined after Bonferroni correction for 18 tests (corresponding to the number of 20° phase bins in the 360° oscillation cycle).

### Behavior assessments and associations with PAC

We evaluated associations of whole brain PAC of theta-gamma, alpha-gamma, and beta-gamma activity with IQ via the DAS-2 and two behavioral domains frequently associated with ASD, social communication (SC) and restricted interests and repetitive behaviors (RRB), as measured by the Social Responsiveness Scale (SRS-2). Because PAC has been shown to change with age (18), we also evaluated PAC association with age and subsequently included age as an independent variable in regression analyses with SC, RRB, and IQ. Associations were run for the full sample (combining ASD and TD groups) and for the ASD group alone. We designated this analysis as an exploratory evaluation of the relationship between PAC and behavior and, therefore, do not correct for multiple comparisons. SPSS by IBM (version 27.0.0.0) was used to run regression analyses.

### Test-retest stability

The above analyses were initially computed on data from timepoint 1 (baseline). Data from timepoint 2 were then used in combination with data from timepoint 1 to evaluate consistency of PAC measures across EEG recording sessions separated by a 6-week period via intraclass correlation coefficients. Only data from participants who participated and contributed good data as determined by HAPPE data quality metrics at both visits were included in this analysis (n = 274), and significance was determined using a two-sided F-test.

## Results

### PAC strength: A priori defined regions of interest and canonical wavebands

First, we evaluated group differences in PAC strength, represented by the modulation index normalized by the surrogate distribution, in frequency pairings corresponding to canonical frequency ranges. This consisted of comparisons between groups in alpha-gamma, theta-gamma, and beta-gamma coupling in anterior, posterior, and all channels. We found significantly increased PAC in ASD when considering theta-gamma coupling over all channels (p < 0.001) and anterior channels (p = 0.001), as well as increased alpha-gamma over all channels that did not survive Bonferroni correction for 9 comparisons (p = 0.045).

### PAC Strength: Data-driven grouping analyses

We next identified significant frequency groupings using a procedure agnostic to canonical frequency ranges typically used in EEG analysis (Figure 2A, 2B). We found significant PAC exclusive to the ASD group in the low frequency phase of around 2-10 Hz signal in frontal channels (FP1, FP2, F7, and F8) (Figure 2D).

**Figure 2:**
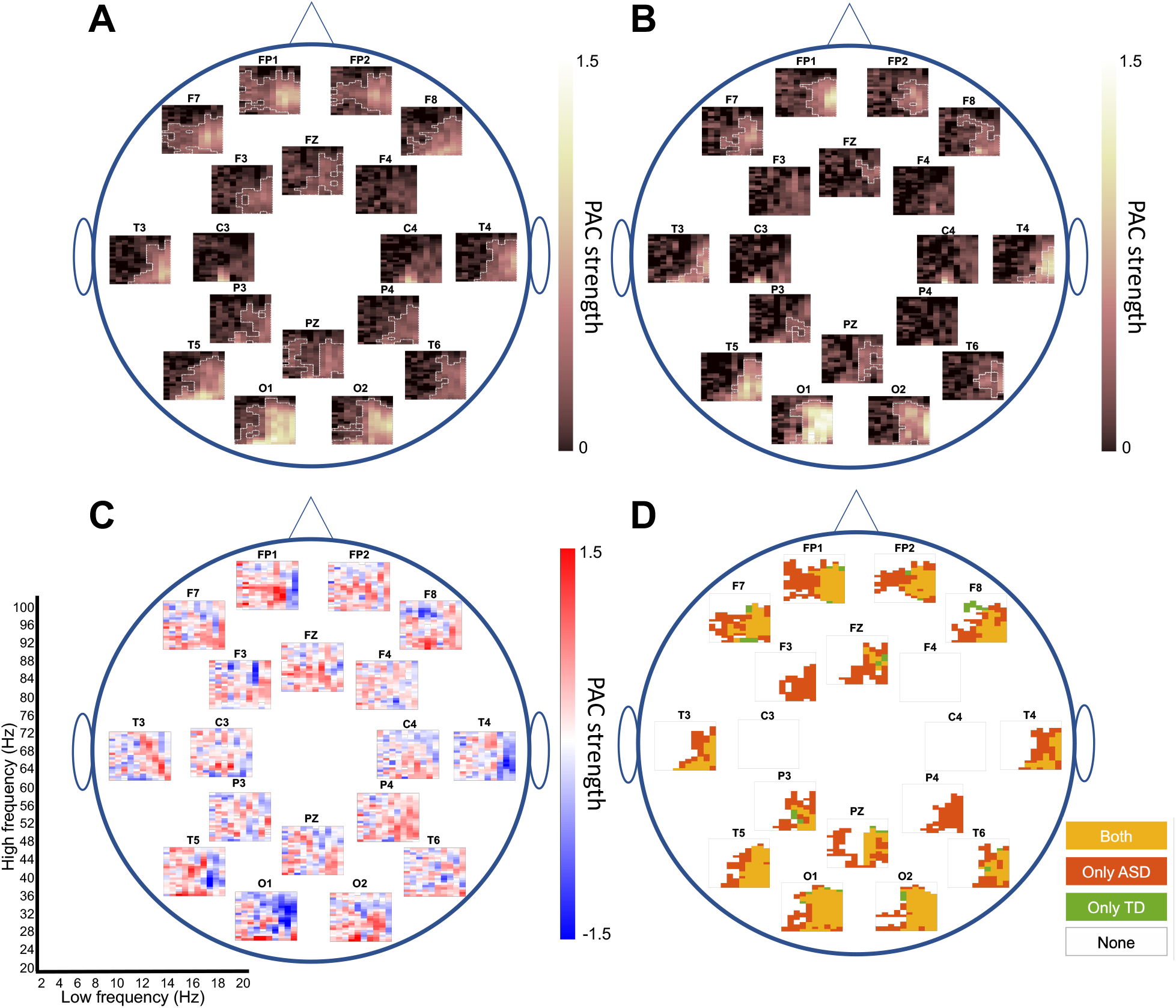
Phase-amplitude coupling strength and significant frequency pairings. Topoplots visualizing PAC strength and frequency grouping information for all channels included in analysis. All subplots within each topoplot share the x- and y-axis of low and high frequency filtered signal, respectively, noted in the lower left corner where each (*x, y*) coordinate on the plot visualizes PAC strength or frequency grouping information for PAC between the corresponding LF *x* and HF *y* signal. A) ASD participant topoplot with normalized PAC strength color-coded using the PAC strength legend increasing from black (low strength) to yellow (high strength). ASD frequency groupings noted with a white outline. B) Topolot same as (A) plotted for the TD group. C) Topoplot visualizing the channel- and frequency-space differences in PAC strength by subtracting TD PAC strength in (B) from ASD PAC strength in (A). Deeper blue frequency pairs correspond to higher PAC strength in TD than ASD, brighter red to higher PAC in ASD than TD, and light colors to areas of marginal PAC strength differences. D) Significant PAC frequency groupings color-coded for ASD and TD groups.

Based on the surrogate distribution for each group, the frequency grouping threshold was 38 and 13 adjacent PAC pairs for ASD and TD, respectively. In order to confirm that size and location of frequency groupings was actually related to diagnosis and not group sample size, we conducted five additional frequency grouping analyses with a randomly selected subset of ASD participants that matched the sample size of the TD group (n=106) (Figure S1). The frequency grouping threshold did decrease from 38 to average 22 pairs, but the general spatial location of frequency groupings nicely corresponded with those determined using the full ASD group, indicating that differences between groups are observed due to participant sample and not size.

We used these data-driven frequency groupings identified with the clustering procedure to further clarify differences between ASD and TD groups. We found that average PAC strength in frequency pairings included in ASD groupings not TD groupings (orange regions in Figure 2D) was stronger in ASD than TD (p < 0.001).

In order to determine whether these group differences were driven by a subset of participants, we next plotted normalized PAC strength for each individual in each of the three a priori defined region of interest and canonical frequency pair combinations where we identified group differences and in channels FP1 and FP2 in the frequency pairs where PAC strength was higher only in the ASD group (Figure 3). Figure 3 show a subset of 14 ASD participants in whom PAC strength was greater than that of all TD participants in at least one of these 4 tests. 8 of these 14 ASD participants consistently showed high PAC strength in 2 or more regions of significant group-level differences. Significant PAC strength group differences were not identified in overlapping significant FP1 and FP2, anterior, and posterior frequency groupings as well as in other canonical frequency pairings.

**Figure 3:**
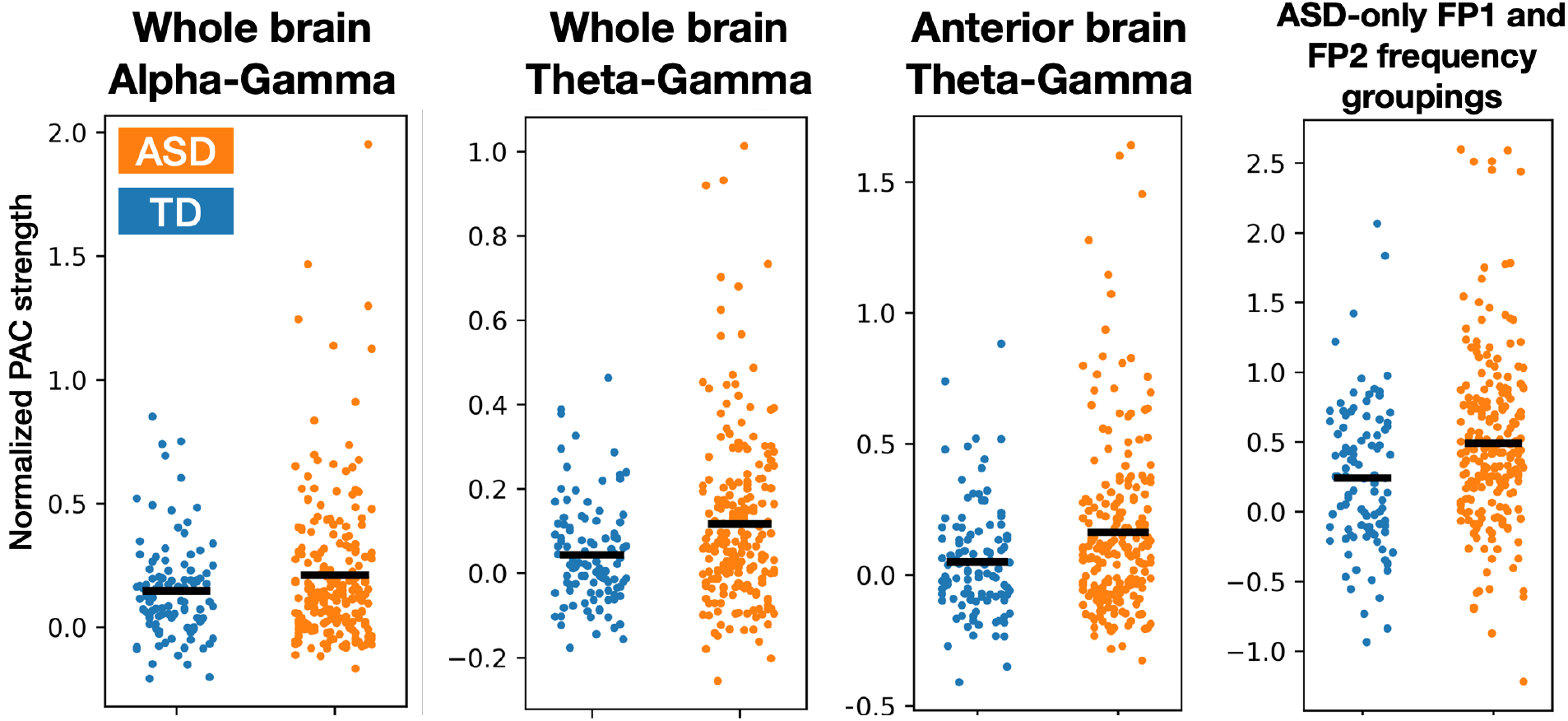
Individual PAC strength Each circle represents the normalized PAC strength value for an individual color coded by diagnosis group (orange for ASD and blue for TD). Group average is indicated by the horizontal black line.

### PAC phase preference

After identifying group differences in PAC strength, we next evaluated what phase of the low frequency signal component was driving these differences. We found significant differences between groups in the theta-gamma phase preference vector averaged for each participant in anterior (p = 0.013) and posterior (p = 0.002) electrodes (Figures 4A and 4B, respectively). We did not identify group differences in PAC phase preference in other regions or canonical frequency pairings.

**Figure 4:**
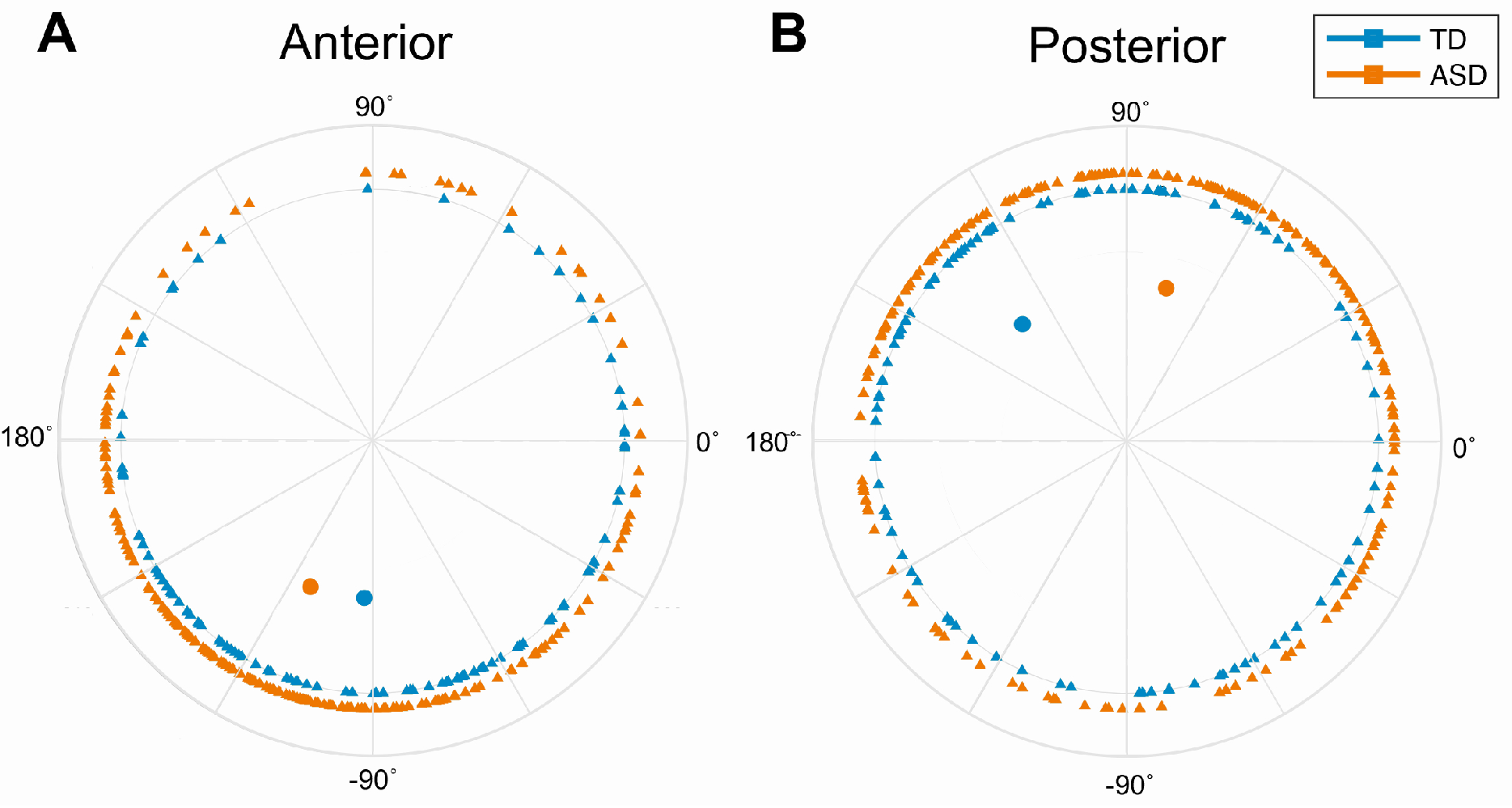
Individual and group average theta-gamma phase preference. Each triangle represents a vector average of phase preference for each frequency pairing falling in the theta-gamma coupling range. Filled circles represent group vector averages of all participants’ average theta-gamma preferences. Color corresponds to outcome group (TD in green and ASD in orange).

Additionally, we evaluated differences in phase bias proportion of theta-gamma coupling in anterior and posterior brain regions. We found that the ASD group was more biased towards - 150° in anterior channels and 30° in posterior channels compared to TD, which corresponded to significant differences in groups 180° from the direction of shift (30° for anterior and -150° for posterior) (Figure 5). This significance corresponded to TD showing less phase shift than ASD, consistent with the finding of stronger PAC in ASD (as strength is directly a result of non-uniform distribution of amplitude in phase bins). Max phase bias proportion of alpha-gamma and beta-gamma coupling did not differ between groups. We are unable to identify outliers using circular statistics given an upper and lower bound for phase preference (e.g., -180° to 180°), which does not exist for z-scored PAC strength.

**Figure 5:**
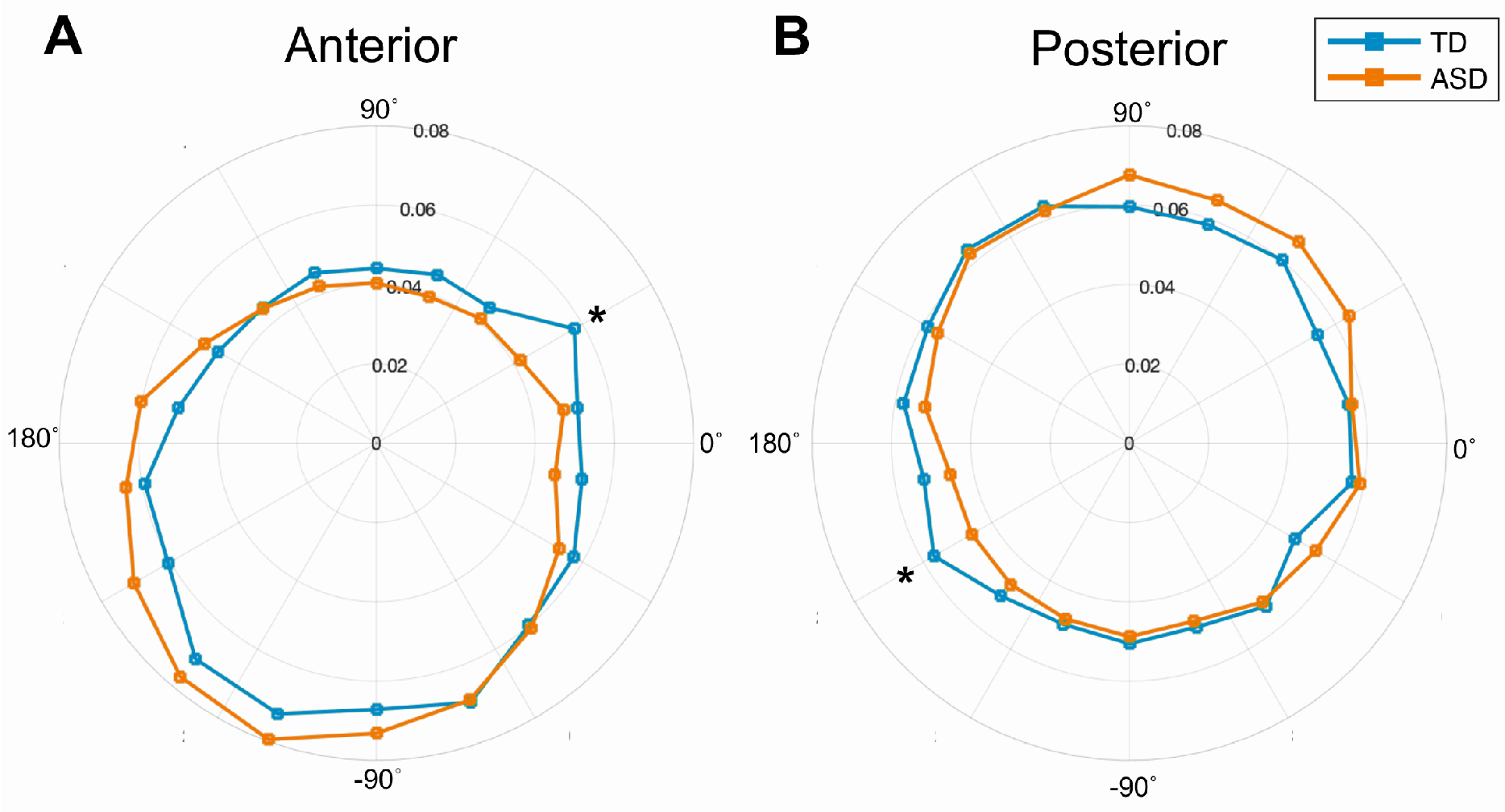
Group differences of phase bias proportion in theta-gamma coupling. Proportion of LF phase bins that contained the maximum HF amplitude for each frequency pairing included in analysis averaged within groups. Color corresponds to group as indicated in the legend. Asterisk indicates significant difference in groups in phase bin (Bonferroni corrected p-value for 18 tests: *p < 0.0028).

### PAC strength and behavioral associations

Age was significantly associated with whole brain theta-gamma and alpha-gamma PAC in the context of all participants and only ASD participants (Table 3). There was a significant association between RRB and SC with theta-gamma PAC as well as SC and IQ with alpha-gamma PAC in the whole group (ASD and TD), but this relationship disappears when only considering ASD participants.

**Table 3:** Linear regression results for three canonical frequency pairs and age, repetitive behavior and restricted interests (RRB), social communication (SC), and full-scale IQ (IQ). Age was included as a control variable for the RRB, SC, and IQ tests. * indicates significance (*p* < 0.05).

| WHOLE GROUP | Brain region & Frequencies | Measure | Beta | *p* value |
| --- | --- | --- | --- | --- |
| Whole brain theta-gamma |  | Age | 0.115 | 0.042* |
|  |  | RRB | 0.166 | 0.003* |
|  |  | SC | 0.185 | 0.001* |
|  |  | IQ | -0.042 | 0.458 |
| Whole brain alpha-gamma |  | Age | 0.223 | <0.001* |
|  |  | RRB | 0.058 | 0.058 |
|  |  | SC | 0.125 | 0.024* |
|  |  | IQ | -0.119 | 0.031* |
| Whole brain beta-gamma |  | Age | 0.037 | 0.515 |
|  |  | RRB | 0.015 | 0.797 |
|  |  | SC | 0.040 | 0.486 |
|  |  | IQ | -0.190 | 0.732 |
| ASD ONLY | Brain region & Frequencies | Measure | Beta | *p* value |
| Whole brain theta-gamma |  | Age | 0.143 | 0.038* |
|  |  | RRB | 0.021 | 0.761 |
|  |  | SC | 0.067 | 0.336 |
|  |  | IQ | 0.043 | 0.536 |
| Whole brain alpha-gamma |  | Age | 0.239 | <0.001* |
|  |  | RRB | 0.031 | 0.649 |
|  |  | SC | 0.063 | 0.360 |
|  |  | IQ | -0.088 | 0.191 |
| Whole brain beta-gamma |  | Age | -0.018 | 0.795 |
|  |  | RRB | -0.025 | 0.724 |
|  |  | SC | 0.065 | 0.352 |
|  |  | IQ | -0.040 | 0.565 |

### PAC strength consistency over recording sessions

To establish performance characteristics of PAC, we computed intraclass correlation coefficients of strength values in participants who contributed data at two sessions separated by 6 weeks (Table 4; Figure S2). Notably, all PAC strength correlations computed for waveband pairs were significant for the ASD group, though inconsistent in the TD group. We discuss the possible impact of ASD outliers on these results in the discussion. There does not seem to be an ICC test adapted to circular statistics, so we are unable to properly evaluate the stability of phase preference over recording sessions.

**Table 4:** ICC statistics for PAC strength in regions and averaged over frequency bands. Values include only participants who participated at T1 and T2 timepoints (n = 174 ASD; n = 88 TD). Bonferroni corrected p-values: * corrected p < 0.05 (raw p < 0.0042); ** corrected p < 0.001 (raw p < 0.000083).

| Region | Waveband pair | ASD ICC [UB LB] | TD ICC [UB LB] |
| --- | --- | --- | --- |
| Whole | Theta-Gamma | 0.47 [0.34, 0.58] ** | 0.26 [0.06, 0.44] |
| Whole | Alpha-Gamma | 0.47 [0.34, 0.58] ** | 0.49 [0.31, 0.63] ** |
| Whole | Beta-Gamma | 0.40 [0.27, 0.52] ** | 0.23 [0.02, 0.42] |
| Anterior | Theta-Gamma | 0.50 [0.38, 0.6] ** | 0.13 [-0.09, 0.33] |
| Anterior | Alpha-Gamma | 0.53 [0.41, 0.63] ** | 0.46 [0.28, 0.61] ** |
| Anterior | Beta-Gamma | 0.31 [0.17, 0.44] * | 0.47 [0.29, 0.62] ** |
| Posterior | Theta-Gamma | 0.40 [0.26, 0.52] ** | 0.28 [0.08, 0.46] * |
| Posterior | Alpha-Gamma | 0.31 [0.17, 0.44] ** | 0.30 [0.09, 0.47] * |
| Posterior | Beta-Gamma | 0.22 [0.08, 0.36] * | 0.23 [0.03, 0.42] |

## Discussion

We found significant differences in PAC measures of resting-state brain activity between children with ASD and TD children. The ASD group showed significant PAC involving the modulation of gamma signal with low frequency (less than 10 Hz) phase in frontal channels and significantly stronger theta-gamma coupling in anterior channels (e.g., where occurrence of significant PAC was identified only in the ASD group) than the TD control group. Additionally, we found shifted phase preference in ASD in both anterior and posterior theta-gamma coupling. Given hypotheses that PAC reflects macroscale interaction between neuronal ensembles (43), the increased PAC strength and shifted phase preference distributed across the cortex in ASD likely reflects altered neural network dynamics. This is of particular importance given the potential for PAC to serve as a marker of the balance between bottom-up and top-down activity, and hence sensory processing and predictive coding, in ASD (17–21). In particular, the theta-gamma frequencies of PAC altered in ASD suggest a preferential shift towards bottom-up processing in this disorder (17).

In general, findings of increased PAC strength in the ASD group were largely driven by a subset of individuals with ASD. Moreover, measures of test-retest reliability of PAC strength suggest that individuals with high PAC strength tend to maintain this high PAC strength across visits (Figure S2). Similarly, individuals with low or medium PAC strength tend to maintain a PAC strength that is in the low-medium range across visits, although their PAC can vary within this low-medium range. This may explain why test-retest reliability of PAC is lower in the TD group (where most individuals have PAC in the low-medium range) than in the ASD group (where some individuals have persistently high PAC) (Figure S2). Overall, this suggests that there is a robust subset of children with ASD who have high PAC.

Nonetheless, we were unable to identify a relationship between these high PAC strength individuals and behavioral assessment scores. Along similar lines, there was a significant positive relationship between RRB, SC, and IQ with PAC strength when considering the entire participant sample, but this did not persist when only considering ASD participants. Of note, a prior study using magnetoencephalography demonstrated a significant relationship between PAC and severity of autism symptoms (44); however, the direction of this relationship varied by brain region and thus may be dampened in our study by the lower spatial resolution of EEG. Another study showed a significant relationship between RRB and alpha-gamma PAC on EEG in children with Phelan McDermid Syndrome, a neurogenetic disorder with high prevalence of ASD (19), although we did not replicate this finding in the present study of children with idiopathic ASD. Notably, there are qualitative group-level differences of RRBs in Phelan McDermid Syndrome compared to idiopathic autism (45). We do find that whole brain alpha-gamma PAC strength increases with age, consistent with prior studies (18,19).

Taken together, these findings raise the possibility that elevated PAC strength may be associated with comorbidities of ASD that were not directly assessed in the present study or may be a marker for an underlying biological process that does not directly correlate with a particular phenotype. For the purposes of biomarker development, it is worth considering the extent to which individuals with high PAC may constitute a biologically if not behaviorally meaningful subgroup. For example, this subgroup might be particularly amenable to neuromodulatory treatments that target bottom-up neural circuit activity, which may not manifest in a single behavioral score.

PAC strength findings contrast with phase preference results, which demonstrate a group-level shift, albeit with substantial overlap between groups, and no discernible subset of individuals driving the findings. Interestingly, the finding of differences in posterior phase preference arises in lieu of differences in posterior PAC strength, indicating that while the posterior PAC may not be notably stronger in one group, there is a shift in low frequency phase peak. Perturbed posterior phase preference is also present in individuals with Phelan-McDermid Syndrome (19), although individual-level phase preference findings were stronger in that study.

Overall, the group-level differences in PAC, particularly in the theta-gamma range, demonstrate potential as a stratification biomarker of ASD and may provide insight into the circuit-level mechanisms underlying some forms of ASD. Several future studies could be considered on the basis of these findings. Back-translation (i.e., examining PAC in animal models) may help elucidate biological underpinnings of altered PAC in ASD and hence inform potential treatment targets. Examination of PAC in related neurodevelopmental disorders may identify a broader spectrum of individuals with altered PAC, who may differ by specific phenotype but share similar circuit function (and thus potentially respond to similar treatments). The present study suggests that PAC may offer clinical utility, and further exploration of this measure (in both directions on the bench-to-bedside spectrum) is necessary to realize this potential.

## Data Availability

All datasets referred to in the manuscript are available via the NIMH Data Archive (identifier: 2288; https://nda.nih.gov/edit_collection.html?id=2288).

## Acknowledgements

Support for this project was provided by the National Institute of Mental Health (U19 MH108206; JCM and R01 MH122428; DŞ). We are grateful to our external advisor board, NIH scientific partners, the FNIH Biomarkers Consortium and all the families and participants who joined with us in this effort. Additional important contributions were provided by members of the ABC-CT consortium. A subset of data from this manuscript were previously presented at the International Society for Autism Research annual meeting, which took place as a virtual event in May 2021.

## Disclosures

SJW has served as a consultant for Janssen Research and Development. GD has served on the scientific advisory boards of Akili Interactive, Hoffmann–La Roche, Janssen Research and Development, LabCorp, Tris Pharma, and Zynerba; she has served as a consultant for Apple, Axial Ventures, Gerson Lehrman Group, Guidepoint Global, and Teva Pharmaceutical; she serves as CEO of DASIO, LLC; she has received book royalties from Guilford, Oxford University Press, and Springer Nature Press; she has developed technology, data, and/or products that have been licensed to Apple or Cryo-Cell International, from which she and Duke University have benefited financially; and she holds a patent (10,912,801) and has patent applications (62,757,234, 25,628,402, and 62,757,226). FS has served as a consultant for BlackThorn Therapeutics, Janssen Research and Development, and Hoffmann–La Roche. JCM has received funding from Janssen Research and Development; he has served as consultant for Blackthorn Therapeutics, BridgeBio, Customer Value Partners, and Determined Health; he has served on the scientific advisory boards of Modern Clinics and Pastorus; and he receives royalties from Guilford, Lambert Press, and Springer. The other authors report no financial relationships with commercial interests.

## Supplemental materials

**Figure S1:**
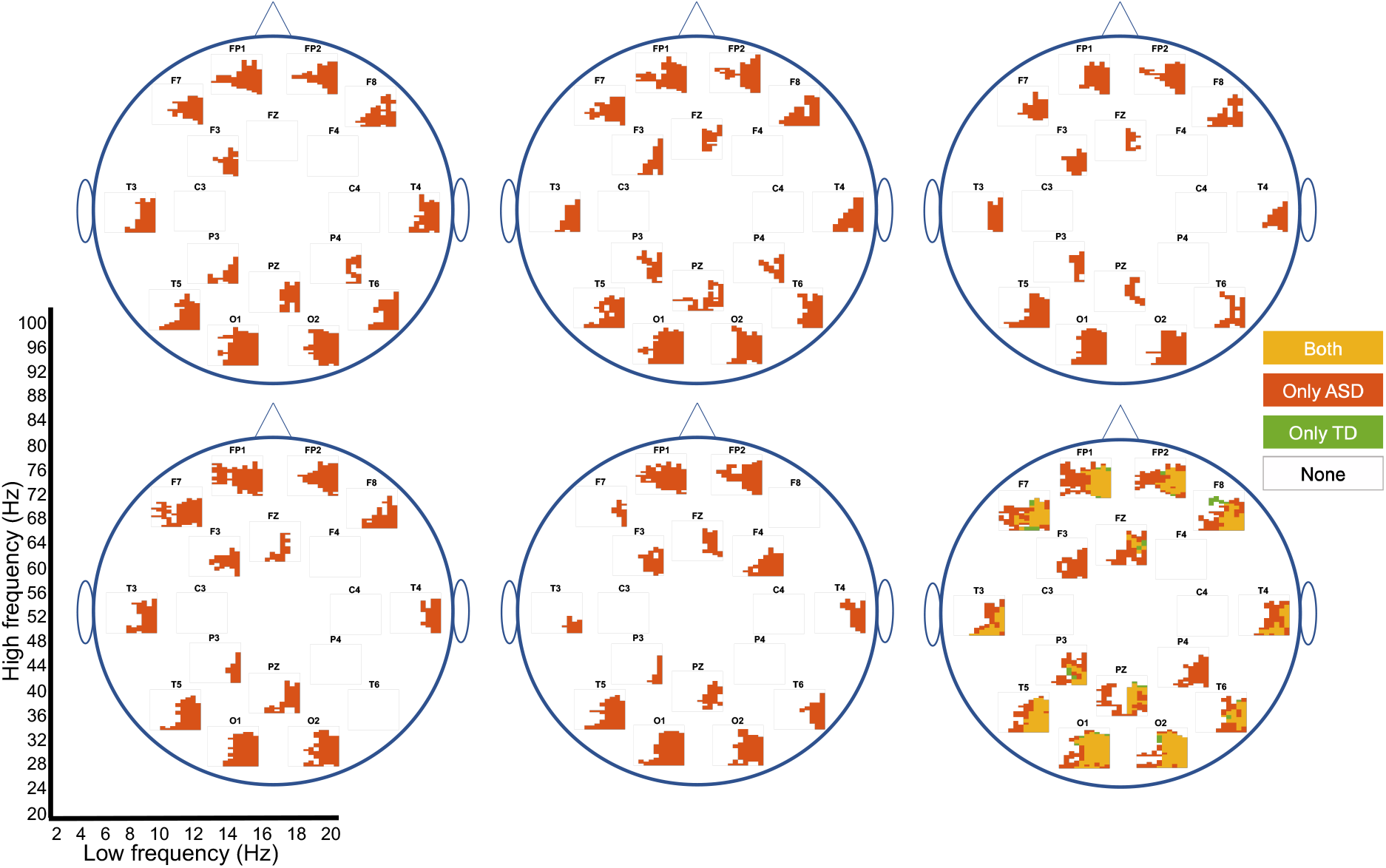
Frequency groupings in smaller sample size of ASD participants. Topoplots visualizing five iterations of significant frequency groupings that arise when 106 ASD participants (to match sample size of TD group) are randomly selected (TD group fully excluded from this analysis). The x- and y-axis noted in the bottom left corner applies to each individual channel plot, and the topoplot in the bottom left corner presents the original frequency groupings for the full ASD group and TD group (same as Figure 2D). Color legend applies to all frequency groupings, where a specific coordinate being colored means that PAC between the corresponding HF and LF frequency belongs to a significant frequency grouping.

**Figure S2:**
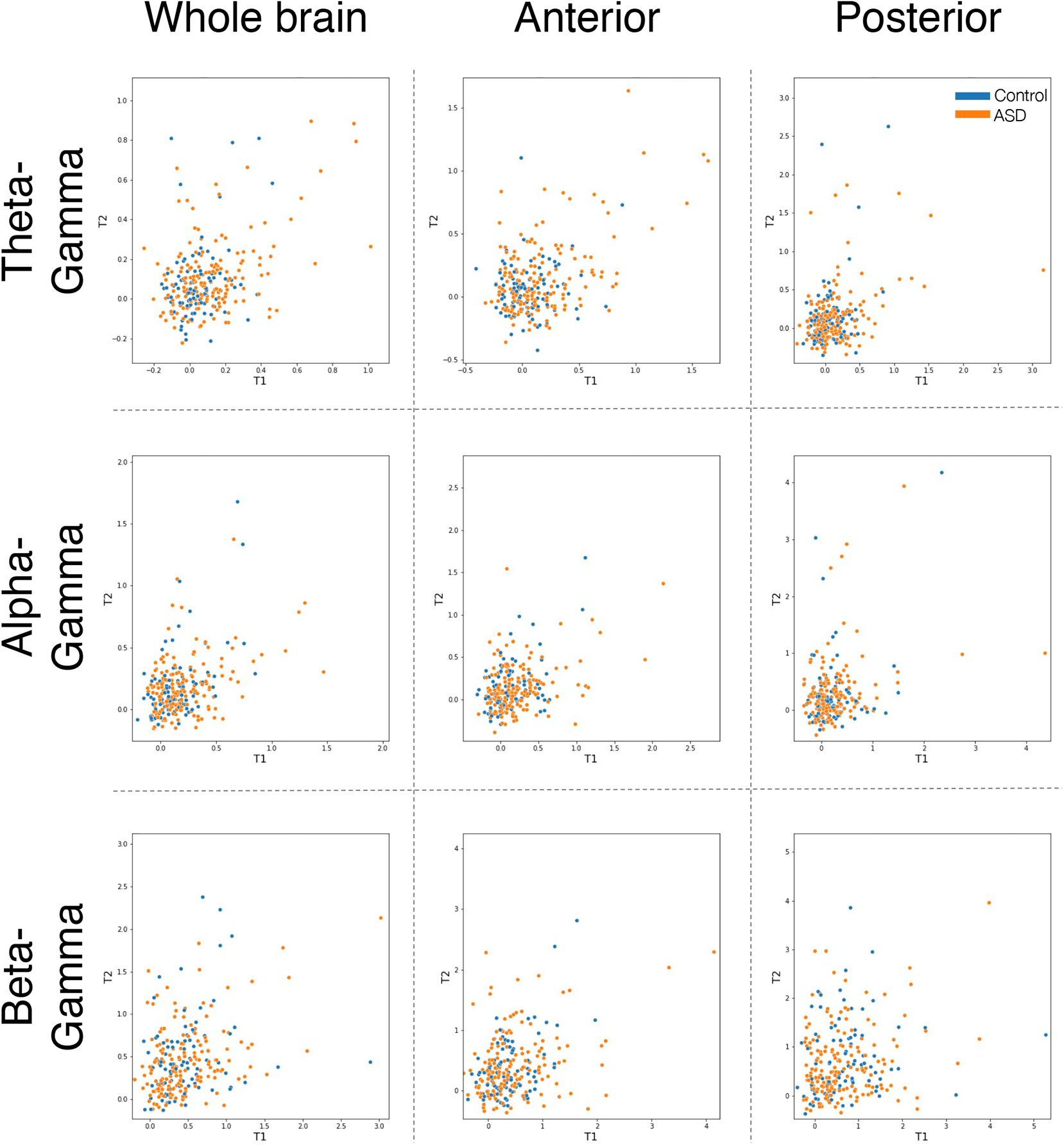
PAC strength consistency across EEG visits. Each plot shows the average PAC strength within the frequency range noted on the y-axis and brain region on the x-axis for each individual participant. Color coded by diagnostic group as indicated by the legend.

## References

1. Mosconi MW, Mohanty S, Greene RK, Cook EH, Vaillancourt DE, Sweeney JA (2015): Feedforward and Feedback Motor Control Abnormalities Implicate Cerebellar Dysfunctions in Autism Spectrum Disorder. Journal of Neuroscience 35: 2015–2025.

2. Khan S, Michmizos K, Tommerdahl M, Ganesan S, Kitzbichler MG, Zetino M, et al. (2015): Somatosensory cortex functional connectivity abnormalities in autism show opposite trends, depending on direction and spatial scale. Brain 138: 1394–1409.

3. Rosenberg A, Patterson JS, Angelaki DE (2015): A computational perspective on autism. Proceedings of the National Academy of Sciences 112: 9158–9165.

4. Gogolla N, LeBlanc JJ, Quast KB, Südhof TC, Fagiolini M, Hensch TK (2009): Common circuit defect of excitatory-inhibitory balance in mouse models of autism. Journal of Neurodevelopmental Disorders 1: 172–181.

5. Peters JM, Taquet M, Vega C, Jeste SS, Fernández IS, Tan J, et al. (2013): Brain functional networks in syndromic and non-syndromic autism: a graph theoretical study of EEG connectivity. BMC Medicine 11: 54.

6. Dinstein I, Heeger DJ, Lorenzi L, Minshew NJ, Malach R, Behrmann M (2012): Unreliable Evoked Responses in Autism. Neuron 75: 981–991.

7. Davis G, Plaisted-Grant K (2015): Low endogenous neural noise in autism. Autism 19: 351– 362.

8. Orekhova E v., Stroganova TA, Nygren G, Tsetlin MM, Posikera IN, Gillberg C, Elam M (2007): Excess of High Frequency Electroencephalogram Oscillations in Boys with Autism. Biological Psychiatry 62: 1022–1029.

9. van Diessen E, Senders J, Jansen FE, Boersma M, Bruining H (2015): Increased power of resting-state gamma oscillations in autism spectrum disorder detected by routine electroencephalography. European Archives of Psychiatry and Clinical Neuroscience 265: 537–540.

10. Yizhar O, Fenno LE, Prigge M, Schneider F, Davidson TJ, O’Shea DJ, et al. (2011): Neocortical excitation/inhibition balance in information processing and social dysfunction. Nature 477: 171–178.

11. Neuhaus E, Lowry SJ, Santhosh M, Kresse A, Edwards LA, Keller J, et al. (2021): Resting state EEG in youth with ASD: age, sex, and relation to phenotype. Journal of Neurodevelopmental Disorders 13: 33.

12. Wang J, Barstein J, Ethridge LE, Mosconi MW, Takarae Y, Sweeney JA (2013): Resting state EEG abnormalities in autism spectrum disorders. Journal of Neurodevelopmental Disorders 5: 24.

13. Webb SJ, Shic F, Murias M, Sugar CA, Naples AJ, Barney E, et al. (2020): Biomarker Acquisition and Quality Control for Multi-Site Studies: The Autism Biomarkers Consortium for Clinical Trials. Frontiers in Integrative Neuroscience 13: 71.

14. Kopell NJ, Gritton HJ, Whittington MA, Kramer MA (2014): Beyond the Connectome: The Dynome. Neuron 83: 1319–1328.

15. Womelsdorf T, Valiante TA, Sahin NT, Miller KJ, Tiesinga P (2014): Dynamic circuit motifs underlying rhythmic gain control, gating and integration. Nature Neuroscience 17: 1031–1039.

16. Sotero RC, Bortel A, Naaman S, Mocanu VM, Kropf P, Villeneuve M, Shmuel A (2015): Laminar Distribution of Phase-Amplitude Coupling of Spontaneous Current Sources and Sinks. Frontiers in Neuroscience 9: 454.

17. Márton CD, Fukushima M, Camalier CR, Schultz SR, Averbeck BB (2019): Signature Patterns for Top-Down and Bottom-Up Information Processing via Cross-Frequency Coupling in Macaque Auditory Cortex. eneuro 6: ENEURO.0467-18.2019.

18. Mariscal MG, Levin AR, Gabard-Durnam LJ, Xie W, Tager-Flusberg H, Nelson CA (2021): EEG Phase-Amplitude Coupling Strength and Phase Preference: Association with Age over the First Three Years after Birth. eneuro 8: ENEURO.0264-20.2021.

19. Mariscal MichaelG, Berry-Kravis E, Buxbaum JD, Ethridge LE, Filip-Dhima R, Foss-Feig JH, et al. (2021): Shifted phase of EEG cross-frequency coupling in individuals with Phelan-McDermid syndrome. Molecular Autism 12: 29.

20. Pellicano E, Burr D (2012): When the world becomes ‘too real’: a Bayesian explanation of autistic perception. Trends in Cognitive Sciences 16: 504–510.

21. Sinha P, Kjelgaard MM, Gandhi TK, Tsourides K, Cardinaux AL, Pantazis D, et al. (2014): Autism as a disorder of prediction. Proceedings of the National Academy of Sciences 111: 15220–15225.

22. Trujillo CA, Gao R, Negraes PD, Gu J, Buchanan J, Preissl S, et al. (2019): Complex Oscillatory Waves Emerging from Cortical Organoids Model Early Human Brain Network Development. Cell Stem Cell 25: 558-569.e7.

23. Port RG, Berman JI, Liu S, Featherstone RE, Roberts TPL, Siegel SJ (2019): Parvalbumin Cell Ablation of NMDA-R1 Leads to Altered Phase, But Not Amplitude, of Gamma-Band Cross-Frequency Coupling. Brain Connectivity 9: 263–272.

24. Takeuchi S, Mima T, Murai R, Shimazu H, Isomura Y, Tsujimoto T (2015): Gamma Oscillations and Their Cross-frequency Coupling in the Primate Hippocampus during Sleep. Sleep 38: 1085–1091.

25. Soplata AE, McCarthy MM, Sherfey J, Lee S, Purdon PL, Brown EN, Kopell N (2017): Thalamocortical control of propofol phase-amplitude coupling ((L. J. Graham, editor)). PLOS Computational Biology 13: e1005879.

26. Jones KT, Johnson EL, Tauxe ZS, Rojas DC (2020): Modulation of auditory gamma-band responses using transcranial electrical stimulation. Journal of Neurophysiology 123: 2504– 2514.

27. Jones W, Klin A (2013): Attention to eyes is present but in decline in 2–6-month-old infants later diagnosed with autism. Nature 504: 427–431.

28. Berman JI, Liu S, Bloy L, Blaskey L, Roberts TPL, Edgar JC (2015): Alpha-to-Gamma Phase-Amplitude Coupling Methods and Application to Autism Spectrum Disorder. Brain Connectivity 5: 80–90.

29. Port RG, Dipiero MA, Ku M, Liu S, Blaskey L, Kuschner ES, et al. (2019): Children with Autism Spectrum Disorder Demonstrate Regionally Specific Altered Resting-State Phase– Amplitude Coupling. Brain Connectivity 9: 425–436.

30. Seymour RA, Rippon G, Gooding-Williams G, Schoffelen JM, Kessler K (2019): Dysregulated oscillatory connectivity in the visual system in autism spectrum disorder. Brain 142: 3294–3305.

31. Khan S, Gramfort A, Shetty NR, Kitzbichler MG, Ganesan S, Moran JM, et al. (2013): Local and long-range functional connectivity is reduced in concert in autism spectrum disorders. Proceedings of the National Academy of Sciences 110: 3107–3112.

32. Mamashli F, Khan S, Bharadwaj H, Losh A, Pawlyszyn SM, Hämäläinen MS, Kenet T (2018): Maturational trajectories of local and long-range functional connectivity in autism during face processing. Human Brain Mapping 39: 4094–4104.

33. Mamashli F, Kozhemiako N, Khan S, Nunes AS, McGuiggan NM, Losh A, et al. (2021): Children with autism spectrum disorder show altered functional connectivity and abnormal maturation trajectories in response to inverted faces. Autism Research 14: 1101–1114.

34. McPartland JC, Bernier RA, Jeste SS, Dawson G, Nelson CA, Chawarska K, et al. (2020): The Autism Biomarkers Consortium for Clinical Trials (ABC-CT): Scientific Context, Study Design, and Progress Toward Biomarker Qualification. Frontiers in Integrative Neuroscience 14. https://doi.org/10.3389/fnint.2020.00016

35. Levin AR, Méndez Leal AS, Gabard-Durnam LJ, O’Leary HM (2018): BEAPP: The Batch Electroencephalography Automated Processing Platform. Frontiers in Neuroscience 12. https://doi.org/10.3389/fnins.2018.00513

36. Gabard-Durnam LJ, Mendez Leal AS, Wilkinson CL, Levin AR (2018): The Harvard Automated Processing Pipeline for Electroencephalography (HAPPE): Standardized Processing Software for Developmental and High-Artifact Data. Frontiers in Neuroscience 12: 97.

37. Mullen T (2012): CleanLine EEG plugin. Neuroimaging Informatics Toolsand Resources Clearinghouse (NITRC).

38. Winkler I, Haufe S, Tangermann M (2011): Automatic Classification of Artifactual ICA-Components for Artifact Removal in EEG Signals. Behavioral and Brain Functions 7: 30.

39. Dupré la Tour T, Tallot L, Grabot L, Doyère V, van Wassenhove V, Grenier Y, Gramfort A (2017): Non-linear auto-regressive models for cross-frequency coupling in neural time series ((F. P. Battaglia, editor)). PLOS Computational Biology 13: e1005893.

40. Tort ABL, Komorowski R, Eichenbaum H, Kopell N (2010): Measuring Phase-Amplitude Coupling Between Neuronal Oscillations of Different Frequencies. Journal of Neurophysiology 104: 1195–1210.

41. Berens P (2009): CircStat: A MATLAB Toolbox for Circular Statistics. Journal of Statistical Software 31: 1–21.

42. Berens P (2009): CircStat: A MATLAB Toolbox for Circular Statistics. Journal of Statistical Software 31. https://doi.org/10.18637/jss.v031.i10

43. Canolty RT, Knight RT (2010): The functional role of cross-frequency coupling. Trends in Cognitive Sciences 14: 506–515.

44. Port RG, Dipiero MA, Ku M, Liu S, Blaskey L, Kuschner ES, et al. (2019): Children with Autism Spectrum Disorder Demonstrate Regionally Specific Altered Resting-State Phase– Amplitude Coupling. Brain Connectivity 9: 425–436.

45. Srivastava S, Condy E, Carmody E, Filip-Dhima R, Kapur K, Bernstein JA, et al. (2021): Parent-reported measure of repetitive behavior in Phelan-McDermid syndrome. Journal of Neurodevelopmental Disorders 13: 1–12.

